# Effectiveness of a school-based multimodal suicide and depression prevention program: cluster-randomized pragmatic trial

**DOI:** 10.64898/2026.04.28.26351983

**Authors:** Mandy Gijzen, Sanne Rasing, Daan Creemers, Rutger Engels, Filip Smit

## Abstract

**Background:** Suicide is one of the leading causes of death among adolescents. Suicidal thoughts and behaviors are significant risk factors for suicide, whilst depressive symptoms are significant risk factors for suicidal thoughts and behaviors. Multimodal school-based interventions that address these risk factors are considered the appropriate approach to suicide prevention.

**Method:** 1,593 adolescents (aged 11-15 years) from 15 high schools in the Netherlands took part in the study, which was designed as a pragmatic cluster-randomized trial. The experimental condition consisted of (1) screening for depressive symptoms and suicidal thoughts and behaviors, followed by referral of those who scored positive; (2) gatekeeper training for school mentors; (3) a serious game to reduce stigma, promote health literacy and improve help seeking behavior; and (4) eight CBT-based sessions for adolescents with elevated depressive symptoms. The control group consisted of screening and the gatekeeper training. Both suicidal thoughts and behaviors and depressive symptoms were assessed at baseline, after 6 months and after 12-months follow-up.

**Results:** Multilevel mixed-model analysis revealed that, over the entire 12-month follow-up period, both suicidal thoughts and behaviors and depressive symptoms increased significantly in the experimental group, but not in the control group. No clinically relevant change was observed in either group.

**Conclusions:** The multimodal stepped-prevention program tested in the present study did not lead to a reduction in suicidal thoughts and behaviors or depressive symptoms. However, it is likely that the adverse impact of COVID-19-related school closures overwhelmed the program’s effectiveness. Furthermore, few high-risk adolescents participated in the CBT-based sessions.

**Key Practitioner Messages:** *What is known?:*

- School-based prevention programs aim to reduce suicidal thoughts, behaviors, and depressive symptoms among adolescents.
- Multi-modal interventions are increasingly promoted, but evidence of their effectiveness is limited under real-world conditions.

*What is new?:*

- The multimodal stepped STORM program did not reduce STBs or depressive symptoms at 12-month follow-up; a small group-level increase was observed in the experimental group.
- Participation in the indicated CBT-based module was low, revealing critical implementation gaps school programs.

*What is significant for clinical practice?:*

- School-based programs should prioritize early identification with active referral, gatekeeper training.
- Active referral to a brief CBT-based intervention for high-risk students need to be implemented in close collaboration with both students and teachers to better assure its uptake

## Introduction

Adolescence is a key period for the onset of suicidal thoughts and behaviours (STBs), which typically emerge between ages 12–16 (Glenn et al., 2017) and peak between 15–29 years (World Health Organisation, 2021; Evans et al., 2005; Nock et al., 2008). Depression is a major co-occurring risk factor that also commonly first appears in adolescence (Goldston et al., 2008; Kerr et al., 2013; Lawrence et al., 2015; Zubrick et al., 2017; Kessler et al., 2005). These overlapping developmental trajectories support integrated prevention of depression and STBs. Meta-analytic evidence indicates that school-based interventions targeting shared risk factors, such as depression, can be as effective as those directly targeting STBs (Gijzen et al., 2022), and that multilevel collaboration may enhance effects (Hegerl et al., 2006).

School-based prevention programs show small but significant effects on both STBs and depressive symptoms, particularly when targeting shared risk factors (Gijzen et al., 2022; Robinson et al., 2018; Werner-Seidler et al., 2021; Walsh et al., 2022). Multimodal approaches may enhance effectiveness (Robinson et al., 2018; Katz et al., 2013), although outcomes depend strongly on implementation quality and contextual fit (Walsh et al., 2023; Kiran et al., 2024). Broader universal school-based programs may indirectly reduce STBs through mechanisms such as emotion regulation and help-seeking (Gijzen et al., 2022; Ayer et al., 2023).

Building on this, the multimodal-stepped depression and suicide prevention program of the Strong Teens and Resilient Minds (STORM) program was developed to initiate and strengthen collaborations between schools, community health services and youth mental health care. The STORM program combines (1) screening of depressive symptoms and suicidal thoughts and behaviors by the municipal health services, followed by in-person assessment and possibly referral to the student’s general practitioner; (2) gatekeeper training for school mentors to create a safety net for students; (3) a universal program for all students using a serious game and a classroom session led by a person with lived experience to reduce stigma and improve health literacy and help seeking; and (4) an indicated prevention program consisting of eight CBT-based skill sessions for adolescents with elevated depressive symptoms.

Despite promising findings, the real-world effectiveness of school-based prevention programs remains uncertain due to implementation challenges such as low participation in indicated components, unclear referral pathways, and variability across schools. In the present study, we evaluated the multimodal stepped STORM program in a cluster randomized controlled trial conducted in secondary schools, as implemented in routine practice with coordination support but without researcher-led delivery of intervention modules. This design provides insight into the performance of a stepped suicide prevention model embedded in school settings while maintaining structured oversight. Schools in the experimental condition implemented the full program, while control schools received screening and gatekeeper training (i.e., enhanced usual care). We examined whether the program reduced suicidal thoughts and behaviors and depressive symptoms over time, and aimed to contribute to the understanding of how integrated school-based prevention can be implemented safely and effectively in real-world settings.

## Methods

### Design

Fifteen high schools participated in the pragmatic cluster randomized trial in two parallel groups. There were four high-school levels involved in the study: (1) pre-lower vocational training, (2) pre-higher vocational training, (3) pre-university training, and (4) combined pre-higher vocational and pre-university training. Schools were randomly allocated to the experimental or control condition stratified for both school level and school size (number of second grade students) by an independent statistician not otherwise involved in the study using permuted blocks randomization using R’s dplyr-package. Schools were randomized rather than students in order to reduce the risk of contamination. Due to the character of the interventions, masking could not be attempted.

The study was approved by the medical ethics committee CMO Regio Arnhem-Nijmegen of the Netherlands (NL61559.091.17) and registered in the Dutch Trial Register for RCT’s (NTR5725). Outcomes are reported in accordance with the CONSORT 2010 statement for reporting parallel group randomized trials (Schulz et al, 2010) and its extension for cluster randomized trials (Campbell et al., 2012).

### Participants

Inclusion criteria for the high school students were: (1) participating in the second grade of the high school (i.e., 8th grade in the US), (2) age between 11 and 15 years, and (3) sufficient fluency in the Dutch language to participate in the interventions and complete the questionnaires. All adolescents from the 15 schools meeting these criteria received information about the study specific to their school. Students were asked to give active consent, while parents gave passive consent and thus had the option to opt-out their child from participation in the study. If either the student did not give consent or the parent opted-out their child, the student did not participate.

### Procedure

A total of 4,505 consenting high-school students were screened for STBs and depression symptoms across 15 high schools during two consecutive study years (October–March 2017/2018 and October–March 2018/2019). Screening was performed by the Municipal Health Service (MHS) as part of the MHS’ routine health survey. Out of 4,505 students, 241 (5%) students screened positive for STBs, and 545 (12%) students for depression symptoms. Students with elevated STBs received a private in-person assessment within 48 hours from the MHS or youth mental health services with referral to the student’s GP if needed and parents were involved. Students with elevated depressive symptoms were encouraged to participate in the CBT-based skills training for depression prevention. These follow-up and referral procedures were repeated after each assessment. All second-year mentors were trained as gatekeepers for STBs to provide an extra safety net.

### Measurements

Students completed questionnaires at baseline (T1), 6-month follow-up (T6), and 12-month follow-up (T12). Adolescents received €5 gift vouchers as a reward for each completed follow-up assessment. The T6 and T12 assessments were partly disrupted by the COVID-19 lockdowns of the schools.

#### Primary outcome: Suicidal thoughts and behaviors

STBs were measured using the Vragenlijst Voor Zelfdoding en Zelfbeschadiging (Questionnaire for Suicide and Self-harm [VOZZ], Huisman & Kerkhof, 2017). It is a self-report questionnaire consisting of 39 items in three clusters: life in general, occurrence of adverse events, suicidal thoughts, and actions in the past week. Only a total score is calculated, with a higher score indicating a higher risk of STBs and a score of 86 or higher indicating that professional help may be required (Kerkhof et al., 2015). The Cronbach’s alpha ranged between .89 and .92.

There is also a short form of the VOZZ, namely the VOZZ-Screen which is mainly used for screening. This consists of the first 10 items of the VOZZ, a score of 39 or higher (i.e., screen-positive) suggesting that help may be needed. Cronbach’s alpha ranged between .80 and .83.

#### Secondary outcome: Depressive symptoms

Depressive symptoms were measured using the Children’s Depression Inventory ([CDI-2], Bodden et al., 2016; Kovacs, 2014). It is a self-report questionnaire consisting of 28 items. Each item comprises of three statements that range in severity from 0 to 2 (e.g. “I feel sad sometimes” = 0, “I often feel sad” = 1, “I feel sad all the time” = 2). Scores range between 0- 56. The CDI-2 was found to have good psychometric properties (Kovacs, 2014). Cronbach’s alpha ranged between .88 and .90.

#### Demographic variables

Demographic variables included gender, birthdate, educational level, and ethnicity of both adolescent and parents (i.e., whether born in the Netherlands or elsewhere).

### Interventions

The STORM program comprises four modules described below. Details about the content of the modules are described more extensively elsewhere (Gijzen et al., 2018).

#### Screening

Students were screened using the VOZZ-Screen and CDI-2 to assess the presence of STBs. Adolescents that scored 39 or higher on the VOZZ-Screen or 2 on item 8 of the CDI-2 were all invited for a personal interview by a professional of either the MHS or mental health services within 48 hours and their parents were informed. These adolescents were still invited to participate in the remainders of the STORM program (i.e., the universal and indicated module).

#### Gatekeepers training

Mentors for second grade students were asked to complete a gatekeepers’ training about the theoretical background of suicidal behaviors. This provided them with tools and confidence to discuss suicidal behaviors with students, and pathways for referring high-risk students to professional care. Prior research found that completion of this program led to increased knowledge of suicidal behaviors, skills and competencies in discussing suicidal behaviors and efficacy in handling suicidal behaviors (Cross et al., 2010; Mo et al., 2018; Wyman et al., 2008).

#### Universal prevention module: Moving Stories

Moving Stories is a serious game-based intervention coupled with a classroom session led by a person with lived experience (Tuijnman et al., 2019). It was based on teen Mental Health First Aid (Hart et al., 2016) and aims to increase students’ knowledge of depression and help strategies while decreasing depression stigma. In the game, students support a fictional peer, Lisa, who shows signs of depression. Students are asked to do five things for Lisa each day and discover which of these actions help Lisa feel better. The students can chat to Lisa using multiple choice conversation options and they receive feedback from Lisa at the end of the day about their actions. After five playing days, a lived experience worker will have a debriefing session with them about how students experienced the game and to take stock of the take-home messages from Moving Stories. Prior research found that it decreased depression stigma and increased mental health literacy among adolescents post-intervention (Tuijnman et al., 2022).

#### Indicated prevention module: At Full Force

At Full Force (in Dutch: Op Volle Kracht; OVK2.0) is based on the Penn Resilience Program (Gillham et al., 2007). It was found to decrease depressive symptoms among Dutch adolescents when used as an indicated prevention (De Jonge-Heesen et al., 2020; Wijnhoven et al. 2014). It is a group intervention consisting of eight weekly 1-hour sessions for three to eight students based around the principles of CBT. Additionally, students complete homework assignments to monitor their mood and promote behavioral activation. More details about the content of the program are described in a protocol paper by (De Jonge-Heesen et al., 2016). The sessions are led by a youth mental health professional from the municipalities where the schools are situated. These professionals were all trained by a psychologist in an extensive 3-day training program.

### Data analyses

#### Sample size and power

Taking into account clustering of students within schools, an ante-hoc power calculation indicated that a sample size of 1290 was needed to detect an expected standardized mean difference d□=□0.38 (or larger) on the central clinical outcome (suicidal behaviors) as statistically significant using at α□=□0.05 (2-tailed) and a power of (1-ß) = 0.80 in a test of independent means (Gijzen et al., 2018). Compensating for 30% dropout would require n = 1844. However, the actual sample size that was obtained was n = 1593 which is still well above n = 1290 when compensating for 30% dropout is not needed in an intention-to-treat (ITT) analysis where missing observations due to dropout are either handled by mixed modeling or multiple imputation as planned.

#### Main analysis

As described in our protocol we used multilevel mixed modeling in Stata 16 (StataCorp, 2019) to take into account the hierarchical structure of the data, where measurements are ‘nested’ in students and students ‘nested’ within schools. Analyses were conducted for both the primary outcome (suicidal thoughts and behaviors) and the secondary outcome (depressive symptoms). To test the effectiveness of the intervention we included the variables condition (experimental vs. control), time (baseline vs. 6-months follow-up and 12-months follow-up), the interaction between condition and time in the fixed part of the model, the outcome measured at baseline as a covariate for baseline adjustment and we included variables that were predictive of dropout. Mixed models give unbiased estimates if dropout is missing completely at random (MCAR). Therefore, we identified whether there were any predictors of outcome and missingness/dropout. Possible predictors of outcome were included as covariates and we evaluated whether this influenced the results. Students and schools were included as level in the random part of the equation.

To aid clinical interpretation, we dichotomized the primary and secondary outcome at their respective clinical cutoff values to identify how many adolescents transitioned from above threshold suicidality and depressive symptoms to below threshold, and vice versa, and to see if these transition rates were different in the intervention and control condition. For suicidality we used a cutoff of 86 or higher to indicate above threshold STBs, and for depressive symptoms we used a cutoff of 14 or higher for above threshold depressive symptoms.

#### Sensitivity analyses

Due to the large attrition rate, the ITT analysis was not only based on mixed modelling, but also additional regression analyses on the wide dataset where missing values were imputed in two different ways: multiple (10-fold) imputation with chained equations (MICE) and multiple (10-fold) imputation with the estimate-maximization (EM) algorithm.

First, predictors of both outcome and missingness were identified. All statistically significant predictors were included in the imputation models. Ten imputed datasets were created (Everitt, 2002). Predictors that were included for suicidal thoughts and behaviors: VOZZ score at screening, two items on perfectionism, three items on worrying, and depression scores at baseline and screening. Predictors for depressive symptoms: VOZZ score at screening and baseline, gender, and CDI-2 score at screening. Regression analyses were used to examine whether condition (experimental versus control) had an effect on the outcome variable (VOZZ suicidal thoughts and behaviors and CDI-2 depressive symptoms) at T12. In order to account for clustering of participants within school locations we used the cluster option as implemented in Stata, which takes into account that observations within schools might be correlated. These sensitivity analyses were conducted for both STBs and depressive symptoms.

## Results

### Recruitment

All adolescents in the second grade of the high school (i.e., 8th grade in the US) at the participating schools were invited to participate if they met the inclusion criteria. In total 1,950 students agreed to participate and returned a completed informed consent form, yet only 1,593 completed at least one assessment. Additional information about the procedure and participant flow can be found in Figure 1.

**Figure 1.**
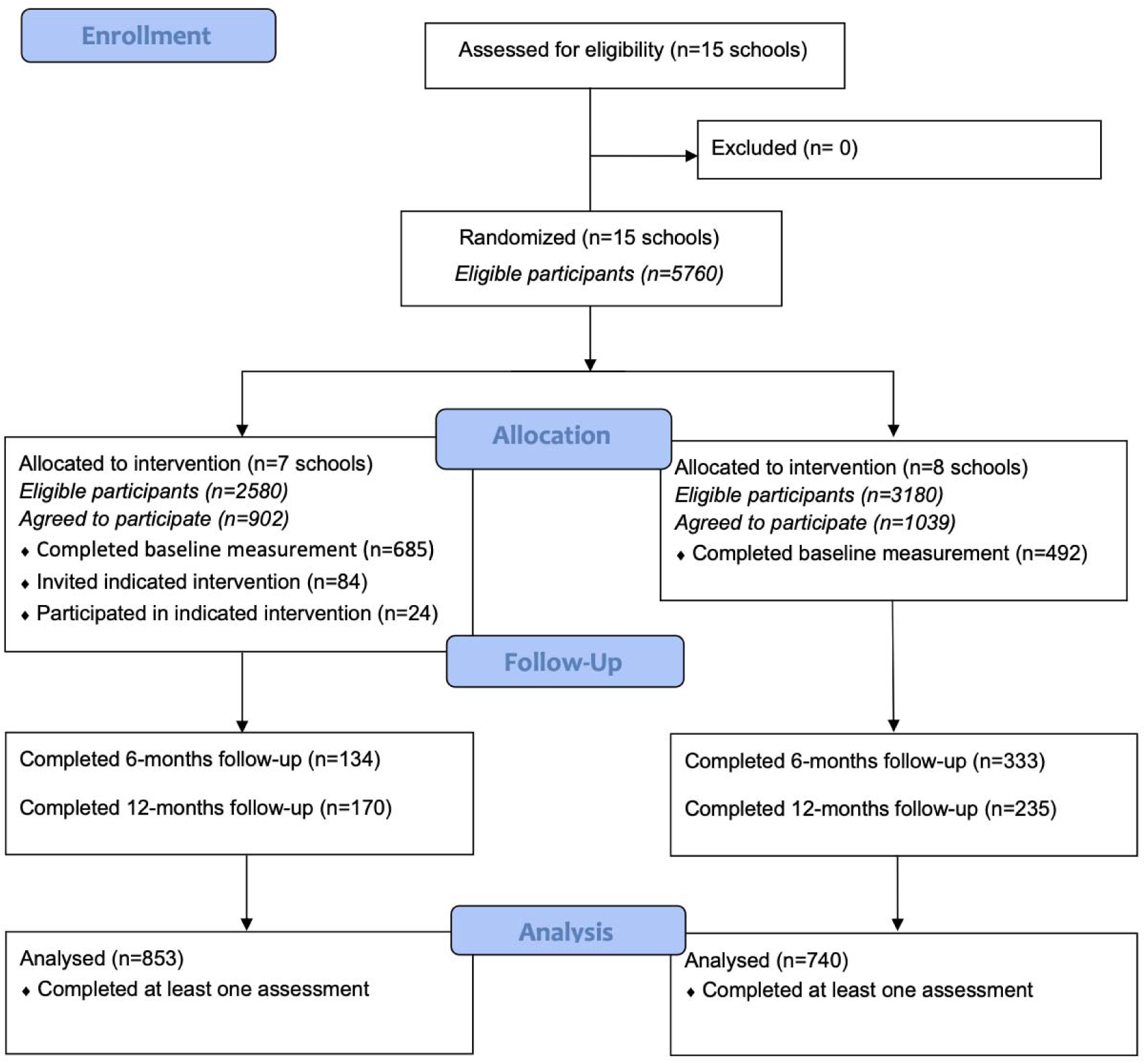
Flow diagram of participants.

### Baseline characteristics

The intention-to-treat sample consisted of 1,593 adolescents of which 853 (53.5%) participants were in the experimental condition. Seven schools were randomized to the experimental condition and eight to the control condition. Table 1 presents the basic demographics and scores on the clinical outcome variables as measured at baseline.

**Table 1.** Baseline characteristics of the participants.

|  | **Total**<br>(N=1593) |  | **Experimental**<br>(N=853) |  | **Control**<br>(N=740) |  |
| --- | --- | --- | --- | --- | --- | --- |
| Age, m (sd) | 13.73 | (1.17) | 13.70 | (1.33) | 13.76 | (.96) |
| Girls, n (%) | 848 | (53.2) | 403 | (54.4) | 445 | (52.2) |
| Suicidal thoughts and behaviors, m (sd) | 61.28 | (15.31) | 61.23 | (14.68) | 61.35 | (16.71) |
| Depressive symptoms, m (sd) | 6.50 | (6.19) | 6.59 | (6.04) | 6.43 | (6.32) |

No baseline imbalance were observed. In other words, cluster randomization resulted in comparable control and experimental group.

### Analysis of drop-out

We conducted logistic regressions to examine attrition at T12 for STBs and depressive symptoms. Across both models, students in the experimental condition were more likely to be lost to follow-up (STBs: *OR* = 1.87, *p* < .001; depressive symptoms: *OR* = 1.86, *p* < .001). Students born in the Netherlands were less likely to drop out (STBs: *OR* = 0.25, *p* = .023; depressive symptoms: *OR* = 0.26, *p* = .028), as were girls (STBs: *OR* = 0.57, *p* < .001; depressive symptoms: *OR* = 0.54, *p* < .001) and students with a higher educational level (STBs: *OR* = 0.38, *p* < .001; depressive symptoms: *OR* = 0.37, *p* < .001). Baseline STBs (*OR* = 1.00, *p* = .240) and depressive symptoms (*OR* = 1.00, *p* = .928) were not associated with attrition.

Mixed models give unbiased estimates if dropout is missing completely at random (MCAR), or when predictors of dropout are included as covariates in the model and then relying on the missing at random (MAR) assumption. Thus, we evaluated if predictors of dropout needed to be included in the model as covariates. The estimates remained nearly identical with or without including predictors of dropout (a difference of less than 0.10 on a scale of 39-195 for STBs and on a scale of 0-56 for depressive symptoms). Therefore, we report only the outcomes of the more parsimonious models that did not include predictors of dropout.

### Participation rates

Of the ITT sample 1,313 students (82.4%) completed the screening for STBs, while 1,314 (82.5%) completed the screening for depressive complaints. A total of 31 students (2.4% of total) scored above-threshold for STBs in the experimental condition (4.5% of experimental group) and 28 (2.1% of total) in the control condition (4.5% of controls). With regards to depressive symptoms, 94 students (7.2% of total) scored above-threshold in the experimental condition (15.8% of experimental group) and 84 (6.4% of total) in the control condition (13.4% of controls). A total of 42 teachers completed the train-the-trainer gatekeeper training after which they ensured that all mentors at their school were trained as gatekeepers.

Furthermore, 982 adolescents downloaded and played the serious game. Adolescents were allowed to participate in the Moving Stories intervention regardless of participation in the research. More information regarding participant rates on Moving Stories can be found elsewhere (Gijzen et al., 2021). Only 24 adolescents completed the OVK2.0 training, this was 29.6% of eligible students with elevated depression scores.

### Outcomes

#### Primary outcome: suicidal thoughts and behaviors

There were no reports of suicides or suicide attempts of participants over the course of the study. Between baseline and 12 months follow-up, we did not find a decrease in suicidal thoughts and behaviors as shown in Figure 2. Suicidal thoughts and behaviors increased in the experimental group compared to the control group and relative to the baseline at T6 (*b* = 2.92, s*e* = 1.27, *z* = 2.31, *p* = .021, 95% CI [.44, 5.40]) and T12 (*b* = 3.42, se = 1.28, *z* = 2.67, *p* = .008, 95% CI [.91, 5.93]) indicating an adverse effect of the intervention at the 6- and 12-months follow-ups. In the control group suicidal thoughts and behaviors remained at baseline level.

**Figure 2.**
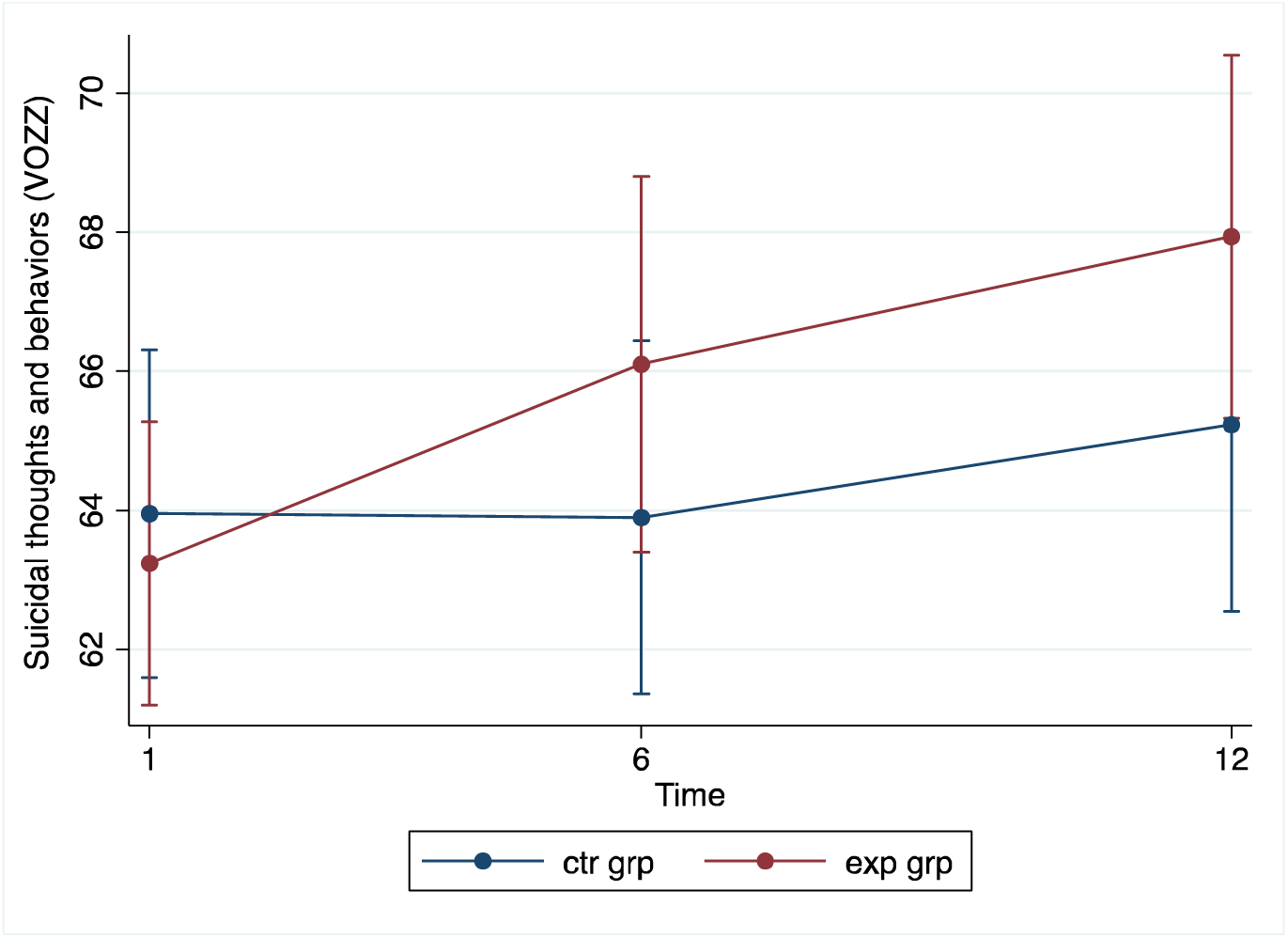
Suicidal thoughts and behaviors by condition over time (with 95% CIs).

**Figure 3.**
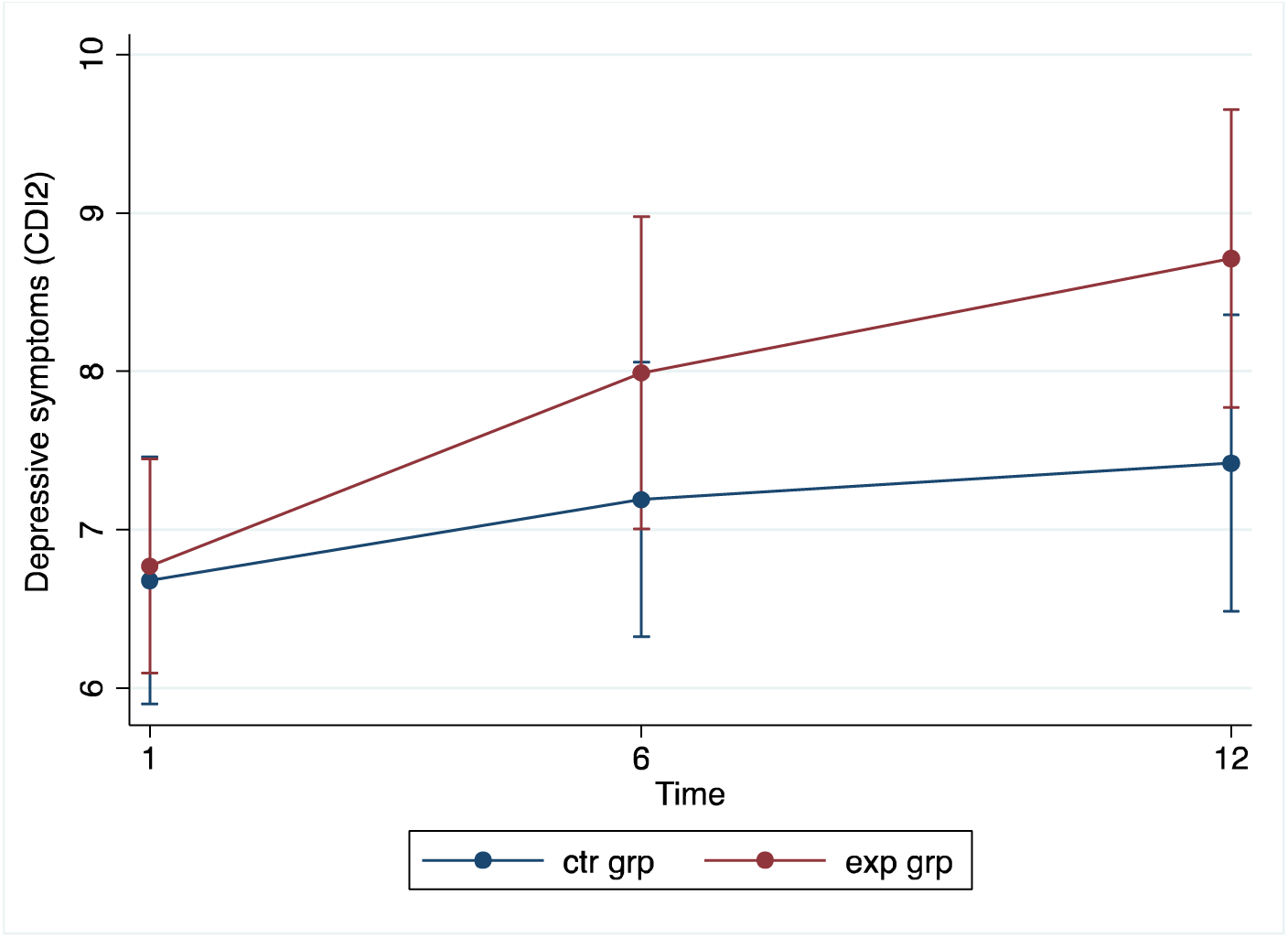
Depressive symptoms by condition over time (with 95% CIs).

Yet, an increase in STBs was noted in both control and the experimental group from baseline to 12-months follow-up. Although the increase of STBs in the experimental group was statistically significant, an increase of about 4 points on a scale of 35-195 cannot be regarded as clinically relevant.

#### Secondary outcome: depressive symptoms

Figure 3 shows that from baseline to 6-month follow-up there was an increase in depressive symptoms, which continued until the 12 months follow-up, and more so in the experimental group than in the control group where depressive symptoms did not change significantly from baseline to follow-up. At the 12-months follow-up this resulted in a significant condition*time interaction effect (*b* = 1.20, *se* =.51, *z* = 2.36, *p* = .018, 95% CI [.20, 2.20]), indicating a significant increase of depressive symptoms in the experimental group compared to the control group.

Yet, an increase in depressive symptoms was noted in both conditions from baseline to 12-months follow-up. Although the increase of depression symptoms in the experimental group was statistically significant, an increase of about 2 points on a scale of 0-56 cannot be regarded as clinically relevant.

For clinical interpretation, suicidal thoughts and behaviors and depressive symptoms were dichotomized at their clinical cut-offs to examine transitions between below- and above-threshold status across groups. Transition probabilities for STBs (VOZZ) and depressive symptoms (CDI-2) are shown in Tables 2 and 3. As expected in a universal prevention sample, most participants remained below threshold, and transition probabilities were comparable across conditions. Transition probabilities from subthreshold (−) to above-threshold (+), and from above-threshold (+) to subthreshold (−), were both approximately 0.30 and did not differ across conditions.

**Table 2.**
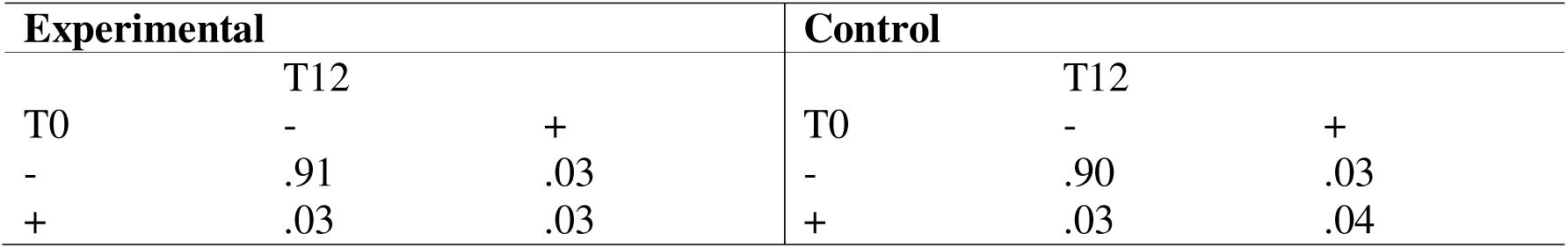
Transition probabilities of suicidal thoughts and behaviors below (–) and above (+) cutoff from T0 to T12 by condition.

| Experimental |  |  | Control |  |  |
| --- | --- | --- | --- | --- | --- |
|  | T12 |  |  | T12 |  |
| T0 | - | + | T0 | - | + |
| - | .91 | .03 | - | .90 | .03 |
| + | .03 | .03 | + | .03 | .04 |

**Table 3.** Transition probabilities of depressive symptoms below (–) and above (+) cutoff from T0 to T12 by condition.

| Experimental |  |  | Control |  |  |
| --- | --- | --- | --- | --- | --- |
|  | T12 |  |  | T12 |  |
| T0 | - | + | T0 | - | + |
| - | .85 | .05 | - | .86 | .04 |
| + | .04 | .06 | + | .04 | .05 |

### Sensitivity analyses

Results from the sensitivity analyses indicated no significant effect of condition on either STBs or depressive symptoms at 12-months assessment.

Results were comparable to the results from the multilevel mixed modelling albeit the sensitivity analyses indicated somewhat lower averages of STBs and depressive symptoms at 12-months follow-up, see Table 4.

**Table 4.** Outcome by main analysis and sensitivity analyses at 12-months follow-up.

| **Outcome (analysis)** | **Experimental group** |  |  | **Control group** |  |  |
| --- | --- | --- | --- | --- | --- | --- |
|  | **Mean** | **95% CI** |  | **Mean** | **95% CI** |  |
| STBs (mixed model) | 67.93 | 65.32 | 70.55 | 65.23 | 62.54 | 67.92 |
| STBs (EM imputation) | 64.85 | 61.60 | 68.11 | 64.19 | 62.75 | 65.63 |
| STBs (MICE imputation) | 65.01 | 61.98 | 68.04 | 63.85 | 61.43 | 66.28 |
| Depressive symptoms (mixed model) | 8.71 | 7.78 | 9.65 | 7.74 | 6.49 | 8.36 |
| Depressive symptoms (EM imputation) | 7.53 | 6.75 | 8.31 | 6.67 | 5.88 | 7.45 |
| Depressive symptoms (MICE imputation) | 7.47 | 6.41 | 8.53 | 6.69 | 5.93 | 7.44 |

## Discussion

The present study evaluated the effectiveness of the STORM multimodal stepped prevention program targeting suicidal thoughts and behaviors (STBs) and depressive symptoms in a high school setting. All participating schools received screening followed by referral to professional care and gatekeeper training, representing enhanced usual care. The experimental condition additionally included a universal (serious game plus a classroom session led by a lived experience worker) and an indicated (OVK 2.0; eight CBT-based group sessions) module. Results therefore reflect the added value of the full multimodal stepped program.

Contrary to our hypotheses, students in the experimental condition showed small but statistically significant increases in STBs and depressive symptoms at 12-month follow-up. However, these effects were not clinically meaningful: scores remained within the non-clinical range and transition probabilities across clinical cut-offs were comparable between conditions. No significant or clinically relevant changes were observed in the control group. COVID-19-related school disruptions may have influenced these outcomes.

### Main findings in context

Evidence on multimodal stepped school-based prevention programs remains limited. Most studies combine screening with either universal or indicated components (Gijzen et al., 2022; Robinson et al., 2018) or target depression or suicidality separately (Gijzen et al., 2022). The EMPATHY program adopted a similar multimodal approach and reported beneficial short-term effects (3-month follow-up) on STBs and depressive symptoms (Silverstone et al., 2015), although the absence of a controlled long-term evaluation limits comparability. Recent meta-analyses suggest effects of school-based prevention programs are limited to post-intervention effects (Robinson et al., 2018), effects on depression diminish beyond six months (Werner-Seidler et al., 2021) and effects beyond 6 months are rarely examined (Gijzen et al., 2022).

The added value of the universal (Moving Stories) and indicated (OVK2.0) modules could not be established. Moving Stories showed limited effects on depressive symptoms and mixed effects on related outcomes such as stigma and helping confidence (Tuijnman et al., 2022). In contrast, OVK2.0 demonstrated effectiveness in reducing depressive symptoms under indicated conditions (De Jonge-Heesen et al., 2020; Wijnhoven et al., 2014, 2016), but not STBs (De Jonge-Heesen et al., 2021). In real-world multimodal implementation, such effects may be further diluted by low uptake and contextual constraints.

At the same time, the observed increase aligns with emerging concerns about potential unintended or iatrogenic effects of universal school-based mental health programs (Guzman-Holst et al., 2025). Their scoping review found that over a third of high-quality studies reported at least one adverse outcome, although such effects are systematically underreported. Risks may arise when universal interventions are insufficiently tailored for adolescents, potentially increasing distress, alienation, or self-stigma (Foulkes & Stringaris, 2023; Foulkes et al., 2024). Proposed mechanisms include emotional overload, mismatch with students’ needs, and negative peer dynamics (Guzman-Holst et al., 2025). Our findings suggest that such adverse trajectories may also occur under real-world conditions. Although causal components cannot be identified, limited uptake of the indicated CBT sessions and mismatch between universal content and students’ needs may have contributed. This is consistent with evidence that only a subset of adolescents perceive CBT-based content as relevant (Bailey et al., 2023), and with broader concerns about applying clinical techniques in universal settings without sufficient developmental contextualization (Foulkes & Stringaris, 2023; Foulkes et al., 2024; Andrews & Foulkes, 2025; Guzman-Holst et al., 2025). Future programs should incorporate monitoring of adverse effects, opt-out options, and adolescent co-design to ensure contextual fit and emotional safety.

Prevention programs, like STORM, are often expected to produce large effects. Even well-established curricula in subjects like math or reading typically produce modest gains (e.g., d ≈ 0.16–0.17; Slavin et al., 2008) yet are widely accepted and implemented due to population-level relevance. In contrast, mental health programs, particularly those targeting STBs, might face pressure to demonstrate strong effects, despite addressing complex and deeply personal issues. Early studies may have contributed to such expectations by reporting inflated effects that do not consistently replicate (Ioannidis, 2005). Accordingly, expectations for universal school-based programs should reflect the complexity of adolescent STBs.

Lastly, COVID-related school lockdowns might have impacted results. The pandemic may negatively affect adolescents’ mental health, increasing depressive symptoms and STBs (Jones et al., 2021) particularly during periods of rising infection rates and restrictions (Reep & Hupkens, 2021; Gezondheidsraad, 2022; Demarest, 2022). Importantly, the timing of school lockdowns relative to assessments differed between conditions (Rijksoverheid, 2022). Students in the control schools that were recruited in our second inclusion year (2018-2019) completed the 12-months follow-up assessments just after the school re-opened, whereas in the experimental group these assessments took place just before, and in the uncertainty of, a lockdown. Such pre-lockdown periods may be particularly stressful, and low-intensity interventions may have limited impact during large-scale stressors (Werner-Seidler et al., 2022). As such, COVID-19 and school lockdowns may have diluted our results, and interpretation should be cautious given the differential impact on experimental and control conditions.

### Strengths and limitations

An important strength was implementation under real-world conditions, enhancing ecological validity and addressing calls for effectiveness research outside controlled environments (Brunwasser & Garber, 2016). A large number of adolescents was reached. Universal screening participation was high (>82%) and implementation included multiple components, such as serious game participation and train-the-trainer gatekeeper sessions, with all mentors at the 15 participating schools ultimately trained as gatekeepers, demonstrating the feasibility of multimodal, school-based prevention program in everyday practice. The 12-months follow-up allowed assessment of the long-term effectiveness and extends beyond most prior studies. The cluster randomized design reduced contamination risk within schools. Lastly, collaborations between key stakeholders in youth mental health were established throughout and after the study. Therefore, there are several effects in both conditions that are beyond the scope of this study.

Several limitations must also be noted. We noted high attrition rates, although sample size was still above the needed sample size without compensating for a 30% dropout. Compensating for dropout is not needed in an intention-to-treat (ITT) analysis where missing observation due to dropout are either handled by mixed modeling or multiple imputation as was planned for the analyses. Missing data was imputed and sensitivity analyses were performed to ensure that results did not rely on chosen method for imputations. Results were highly comparable across imputation methods, indicating that the findings are robust. In addition, the study design precludes conclusions about individual components. Another methodological consideration is the use of the VOZZ as the primary measure of STBs. While it captures cognitive, emotional, and situational aspects of suicidality, it also includes items reflecting general distress and adverse life events, potentially reducing specificity for suicidal ideation or intent.

### Implications

True-to-model implementation remains essential for prevention effectiveness. Exposure to the universal procedure could not be determined and only 24 students (29.6%) participated in the indicated module. While similar to other indicated depression prevention studies (e.g., De Jonge-Heesen et al., 2020: participation rate 27.7%; Van den Heuvel et al., 2021: 14%), this reflects a major implementation gap. We were not able to test the multimodal stepped program as intended, e.g. the added value of both a universal and indicated prevention module. Rather, the participation rates of the indicated intervention module (OVK2.0) were too low to expect an effect of this intervention on STBs or depressive symptoms. This is consistent with evidence from an individual patient data meta-analysis showing that CBT-based prevention requires relatively large samples (number needed to treat = 19 at six months; Rohde et al., 2018), implying that larger sample sizes are needed to really impact depression among adolescents. Low uptake might have diluted the overall effect of the STORM program.

Low participation in indicated modules is common as adolescents with subthreshold symptoms rarely seek help (Cuijpers et al., 2010). Yet, people who completed a prevention program are less likely to develop a depression (Cuijpers et al., 2008). Therefore, both researchers and schools consider how to close the gap between eligibility and participation (Cuijpers et al., 2013) and identify factors that facilitate true-to-model implementation.

## Conclusion

The multimodal stepped-prevention program STORM did not reduce STBs or depressive symptoms. Small significant increases were observed in the experimental group at 12-month follow-up, though changes were not clinically meaningful. COVID-19-related school disruptions may have contributed, but the findings also raise questions about the added value of the universal (Moving Stories) and indicated (OVK2.0) components. Low participation in OVK2.0 highlights a key implementation gap underscoring even well-designed interventions fail if follow-up care is underutilized.

Moving forward, STORM should prioritize screening with active referral and gatekeeper training, alongside improving engagement and adapting the universal and indicated modules. Enhancing uptake to address the implementation gap, incorporating adolescent co-design, and aligning interventions with the social and contextual realities of young people will be critical to ensuring effective and scalable school-based prevention.

## Acknowledgements

We would like to acknowledge Rian van den Boogaart (project manager at GGZ Oost Brabant) for her contribution to design and practicability of the study, and Anouk Tuijnman (Trimbos Institute) and IJsfontein for their contribution in developing Moving Stories. We are also grateful to the collaborating schools (Alfrinkcollege, Carrolus Borromeus College, Commanderijcollege, Dr. Knippenbergcollege, Jan van Brabant College, Hub van Doorne, Peellandcollege, St. Willibrord Gymnasium, Strabrecht College, Vakcollege Helmond, and Varendonck College), the health professionals of the Municipal Health Services “Brabant-Zuidoost”, mental health professionals of GGZ Oost Brabant, Marianne van Bakel for training the experiential experts and the experiential experts we trained for making this research possible.

## Ethics approval and consent to participate

The medical ethics committee CMO Region Arnhem-Nijmegen in The Netherlands approved this study (NL61599.091.17). Written informed consent from adolescents and parents was obtained.

## Data availability

The data that support the findings of this study are not publicly available due to ethical and privacy restrictions. The first author (MG), who conducted the analyses during the study period, had access to the raw data at that time. Access to the data is currently restricted and is held by GGZ Oost Brabant. Requests for access may be directed to GGZ Oost Brabant and are subject to approval by the relevant data governance and ethics committees.

## Authors’ contribution

MG was responsible for data collection, data analysis, and for reporting the study results. FS was involved in the data analysis. SR, FS, DC, and RE read the manuscript and provided suggestions for improvement. SR, DC, FS and RE were also supervisors and grant applicants. All authors have read and approved the final manuscript.

## Trial registration

The study is registered in the Dutch Trial Register (NTR6622).

## Ethical approval

CMO Regio Arnhem-Nijmegen of the Netherlands (NL61559.091.17)

